# Characterising trajectories of social transition in a cohort of gender diverse Australian children and adolescents

**DOI:** 10.1101/2025.10.26.25338420

**Authors:** Will Conabere, Pip Buckingham, Sarah Martin, Ken C. Pang, Michelle Tollit

## Abstract

**Background:** Social transition, defined as all outward changes in gender expression to align with and affirm one’s gender identity, has been richly described in qualitative studies, but quantitative research often lacks the detail required to capture its full complexity. To date, no studies have quantitatively examined how this process varies in young people over time.

**Methods:** Longitudinal data were obtained from 234 trans and gender diverse Australian young people (ages 8-17) attending a specialist paediatric gender service across at least two annual waves from 2017 to 2024. Social transition was operationalised using three practices (name, pronouns, and appearance) across five contexts (home, school, online, with friends, and extended family). For each practice, we captured the contexts in which a change had been enacted or was desired in the future.

**Findings:** Most young people reported changes to their enacted social transition (73.9%) and adjustments in their social transition goals (60.7%) over time. Grouping of participants with similar social transition trajectories identified that most (78.6%) met their goals by the final wave, while many of those who had not were still progressing toward them (50.0%). Progress varied by gender, with trans boys significantly more likely than trans girls to meet their social transition goals.

**Conclusion:** In our longitudinal clinical cohort of gender diverse young people, social transition encompassed diverse practices, contexts, and goals, which evolved over time. Despite this variability, clusters of trajectories emerged, indicating that groups of young people can follow similar patterns, while maintaining unique experiences.

## Introduction

An estimated 1-3% of children and adolescents identify as transgender (Coleman et al., 2022) and, for many of these young people, social transition represents an important means of affirming their gender. Social transition is defined across the literature in many ways (Conabere et al., 2025), but is commonly understood to comprise all outward changes in gender expression that trans individuals undertake to affirm their gender identity. This includes, for example, gender-aligned changes to name, pronouns, and/or presentation (e.g. clothing, hairstyle, chest binding). As such, this process can be highly variable, reflecting the diverse ways young people enact social transition practices across different contexts or to varying extents (Buckingham et al., 2025). For example, some trans young people may avoid all social transition practices at school, or with their extended family, but may feel comfortable using their affirmed name and pronouns with close friends and family. Moreover, social transition is also a dynamic process that can progress, persist, or pivot in line with evolving transition goals.

Variability in navigating social transition has been well-documented in qualitative research, capturing the different practices and contexts young people seek, as well as the positive impacts these changes can bring (de Castro Peraza et al., 2023; Horton, 2022; Wong & Drake, 2017). In contrast, much of the quantitative literature reduces this heterogeneity of experience to limited categories, such as “socially transitioned” or “not socially transitioned”. (Conabere et al., 2025; Horton, 2024). These simple approaches cannot adequately capture young people’s experiences. For example, name change has been used in some past quantitative studies to categorise a participant as “socially transitioned”, yet many trans young people don’t have a desire to change their name (Horton, 2024; Russell et al., 2018). The same applies to all other aspects of social transition, where practices expected by researchers may not be meaningful or desirable to all participants (Pollitt et al., 2019). Further, some young people may wish to make social transition changes in certain contexts but intentionally avoid doing so in others (Olson, 2016), due to safety concerns, fear of transphobia, or simply personal preference. These qualitative studies, and some more recent quantitative approaches (Buckingham et al., 2025), highlight the importance of measuring individual-level differences of social transition, particularly whether non-enacted changes are desired but unmet, or not desired at all.

The process of social transition has been subject to increased public debate and scrutiny in recent years, much of which has stemmed from the publication of the Cass Review of NHS England gender services for children and young people (Cass, 2024). The report recommends a “cautious approach” to social transition, involving clinician oversight and advising that such changes be delayed until after puberty (Cass, 2024). Although the Cass Review does not explicitly define social transition as a binary construct, its framing and interpretation of the literature suggest a fixed, linear, and uniform process that risks inevitably progressing young people toward medical intervention. As outlined earlier, however, social transition is more accurately understood as a dynamic and highly individual process, shaped by both personal goals and levels of safety in varying social contexts. Many quantitative studies aiming to describe social transition or examine associated outcomes (e.g. mental health) omit these critical factors, and as such, their findings have often been overgeneralised. In reality, non-significant mental health differences between socially transitioned and non-socially transitioned youth (Hall et al., 2024) likely reflect (1) societal consequences of social transition (e.g. facing increased transphobia, contributing to poorer mental health outcomes), (2) the level of social support received during their social transition (e.g. those supported in their identities tend to have more favourable outcomes), and (3) whether a young person wishes to enact a given change (e.g. young people who do not wish to change their name may be mischaracterised in research as non-socially transitioned) (Durwood et al., 2021; McGuire et al., 2010; Trujillo et al., 2017; Turban et al., 2021a; Simons et al., 2013).

Another limitation in the existing social transition literature is the predominance of cross-sectional studies, capturing only a single moment of an inherently dynamic process. Longitudinal studies in other areas of transgender health have been important in understanding fluidity of gender identity (Durwood et al., 2022; Katz-Wise et al., 2024) and trajectories following medical treatment (Brik et al., 2020; van der Loos et al., 2023). Consistent with this, the present study tracks participants’ social transitions over time to capture the complexity of this process and to identify patterns or subgroups that cannot be detected in cross-sectional research. More specifically, this study utilises rich longitudinal quantitative data on social transition among young people aged 8-17 years accessing gender affirming care in Victoria, Australia, and aims to:

- Describe the social transition trajectories of these young people, including which practices are enacted, the contexts in which they may or may not occur, and desire to enact future changes;
- Identify common trajectory patterns that are shared between groups of participants who have enacted similar social transition practices and expressed similar social transition goals over time; and
- Characterise the demographic profiles associated with each distinct trajectory.

## Methods

### Study Design

This longitudinal descriptive study is reported in accordance with the STrengthening the Reporting of OBservational studies in Epidemiology (STROBE) statement (von Elm et al., 2007).

### Setting and Participants

This study draws on data from the Trans20 cohort (Tollit et al., 2019), consisting of 618 young people recruited from 2017 to 2020 through the Royal Children’s Hospital Gender Service (RCHGS) in Victoria, Australia. RCHGS is a multidisciplinary specialist paediatric gender clinic that, as part of standard clinical care, administers annual surveys to young people and their nominated parent or caregiver. The Trans20 study was approved by the Royal Children’s Hospital Research Ethics Committee (#36323).

The first survey was administered to RCHGS patients approximately 1 month before an initial triage appointment (these cross-sectional social transition data are reported elsewhere, see: Buckingham et al., 2025). The next survey was administered just prior to the young person’s first appointment with the multidisciplinary assessment clinic (MDAC), which consists of a series of 6-8 appointments over ∼6 months with mental health clinicians and paediatricians or endocrinologists, where clinicians assess suitability for medical affirmation. Subsequent surveys were sent out on an annual basis. In this study, the survey completed prior to the first MDAC appointment were used as the baseline to minimise attrition. Participants were eligible for inclusion in this study if they were part of the Trans20 cohort, responded to at least one question in the social transition section in the pre-MDAC survey and at least one subsequent survey, and were aged 8 years or older (social transition measures were not included in surveys for those <8 years). The sample selection process is outlined in Figure 1.

**Figure 1:**
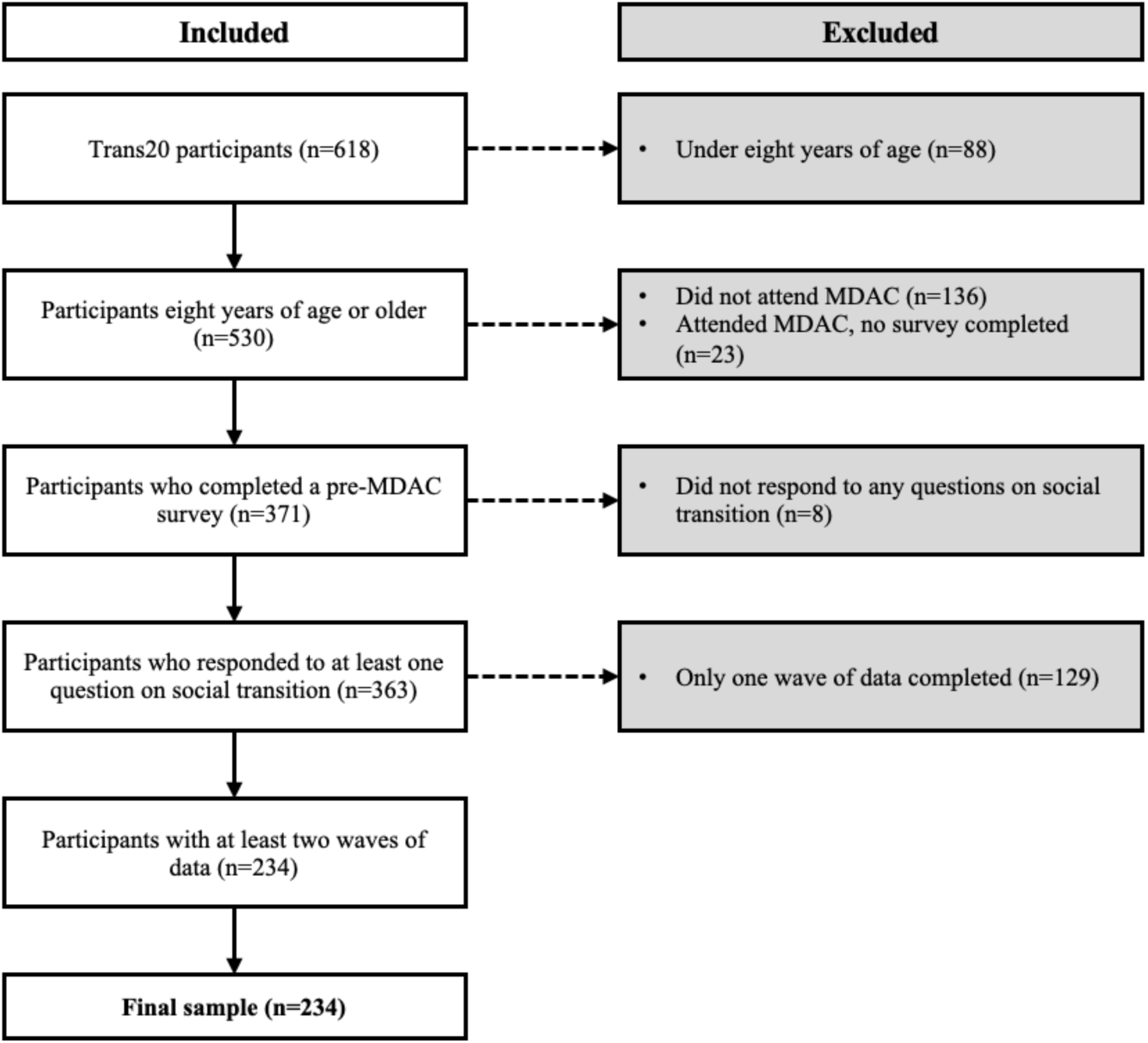
Participant inclusion and exclusion criteria flowchart. Trans20 participants are in the broader Trans20 study (see reference in text); MDAC indicates the Multidisciplinary Assessment Clinic at the Royal Children’s Hospital Gender Service, where ongoing care is provided for the young person.

### Data sources

Data were collected in LimeSurvey (LimeSurvey GmbH, n.d.) from Trans20 surveys completed by the young person and their parent across two or more consecutive waves between 2017-2024, commencing at first MDAC visit and then approximately 12 monthly. The surveys collect information about gender identity, affirmation, health, and wellbeing. Survey links were emailed to parents, who completed their own questionnaire and then provided the young person’s survey link to their child. All participants were informed that their responses would not affect their access to care. Data were also drawn from the patient electronic medical record (EMR), which collects demographic and treatment information. Demographic variables including participant age, country of birth, language spoken at home, socioeconomic status, rurality, school attendance frequency, and identification as Aboriginal and/or Torres Strait Islander were determined from an audit of all three data sources. Gender identity was collected in young person surveys and final categories were derived using coding rules described previously (Blacklock et al., 2021). To assess socioeconomic status, we linked residential postcodes from the EMR with the Index of Relative Socioeconomic Advantage and Disadvantage (IRSAD; Australian Bureau of Statistics, 2021). To assess rurality, we linked these residential postcodes with the Modified Monash Model (MMM) rurality index (Australian Government Department of Health and Aged Care, 2024). Data on social transition were captured exclusively from the young person’s surveys.

### Social transition constructs and measures

Social transition is operationalised in this study with multiple constructs and measures. First, we examined three *practices* (name, pronouns, and presentation), with each assessed across five *contexts* (home, school, with friends, with extended family and online – the latter only in those aged >11 years in accordance with the legal age of social media use in Australia at the time Trans20 commenced). Response options (Table 1) indicated whether a practice was enacted (“Done”), desired (“One day”), or, not desired (“I don’t want to”) in each context, respectively. This approach generated 15 datapoints per participant to capture their social transition at each timepoint.

**Table 1:** Social transition practices and contexts assessed in this study, drawn from the Trans20 questionnaires.

| Construct measured | Response options |
| --- | --- |
| Change of name: |  |
| At home | Individually for each location:<br>▪ ‘Done’<br>▪ ‘One day’<br>▪ ‘I don’t want to’<br>▪ ‘I don’t know’<br>▪ ‘Not applicable’ |
| At school |  |
| Online |  |
| With friends |  |
| With extended family |  |
| Change of pronouns: |  |
| At home | Individually for each location:<br>▪ ‘Done’<br>▪ ‘One day’<br>▪ ‘I don’t want to’<br>▪ ‘I don’t know’<br>▪ ‘Not applicable’ |
| At school |  |
| Online |  |
| With friends |  |
| With extended family |  |
| Change of presentation: |  |
| At home | Individually for each location:<br>▪ ‘Done’<br>▪ ‘One day’<br>▪ ‘I don’t want to’<br>▪ ‘I don’t know’<br>▪ ‘Not applicable’ |
| At school |  |
| Online |  |
| With friends |  |
| With extended family |  |

### Operationalisation of social transition trajectories

To characterise each participant’s trajectory of social transition, we derived three variables for each of the three measured practices (name, pronouns, presentation): (i) completeness of desired social transition, (ii) direction of enactment between waves, and (iii) change to social transition goals. Details of the calculation for each variable are provided below, and an example of these calculations is illustrated in Supplementary Figure 1.

#### Trajectory variable one: Completeness of desired social transition

Completeness was defined as the proportion of desired contexts in which a participant had enacted each social transition practice. A separate completeness metric was calculated for each practice (name, pronouns, and presentation), giving each participant one value per practice at each wave. The numerator counted the number of contexts where the practice had been enacted (e.g. a participant reporting a pronoun change at home, with friends, and online = 3), and the denominator counted the number of contexts where a change had either been enacted or was desired (e.g. the same participant desired a change in one additional context, giving a total of 4). Contexts where a change was neither enacted nor desired were excluded from both the numerator and denominator for that individual. Responses of “I don’t know” or “Non applicable” were coded as missing to avoid misrepresenting the participant’s intentions. Using this calculation, the completeness metric quantifies the extent to which each participant’s desired social transition had been enacted during that wave for each practice (e.g. in the example above, pronouns completeness = 3/4, or 75%).

Completeness metrics were analysed as a continuous measure over time, to show progression toward desired social transition. However, to aid interpretation and grouping, we categorised participants into the following groups:

- **Meeting social transition goals** at all waves (all completion metrics = 100%);
- **Not meeting social transition goals** at any wave (all completion metrics ≠ 100%);
- Had not initially, but **progressed to meeting social transition goals** by the final wave (initial completion metric ≠ 100%; final completion metric = 100%);
- Had initially but **are no longer meeting social transition goals** by the final wave (initial completion metric = 100%; final completion metric ≠ 100%).

#### Trajectory variable two: Changes to enacted social transition

Changes in enacted social transition over time were derived from the numerator of the completeness metric. For each practice, the number of contexts where a change had been enacted was compared between waves (Waves 1 vs 2, 2 vs 3, and 1 vs 3). Participants were then categorised as:

- **Enacting social transition changes**: changes occurred in only one direction (increase) since baseline.
- **Removing social transition changes**: previously enacted changes removed in only one direction (decrease) since baseline.
- **No change**: no changes introduced or removed since baseline.
- **Fluctuations**: changes occurring in varying directions between waves.

#### Trajectory variable three: Adjustment of desired social transition

Changes in desired social transition were derived from the denominator of the completeness metric. Comparisons between waves were used to classify participants as:

- **No change** to social transition goals.
- **Increased social transition goals**: new contexts became desired.
- **Decreased social transition goals**: previously desired contexts no longer desired.

## Data analysis

### Demographic characteristics and comparisons across waves

Descriptive statistics were used to summarise demographic variables for the baseline (pre-MDAC) sample (n=371), the subset included in the trajectories analysis (i.e. individuals with at least two waves of social transition data, n=234) and those lost to attrition (i.e. only one wave completed, n=137). Frequency (n) and column-wise percentage (%) were reported for categorical variables, while mean and standard deviation (SD) were used for continuous variables. Differences in categorical variables were assessed using Chi-square tests of independence for each demographic variable, and differences in continuous variables were assessed using a one-way ANOVA. Significant differences between groups were investigated using pairwise comparisons, using proportions and their 95% confidence intervals for Chi Square results, and Tukey’s test for ANOVA results. Statistical significance was set at p<.05 for all analyses.

### Individual trajectories

To examine individual patterns of social transition over time, individual trajectories were visualised using heatmaps. Heatmaps illustrated changes in the completeness metric across the three waves, showing how each participant enacted, persisted with, or removed, desired practices over time. Each practice (name, pronouns, and presentation) is displayed separately to allow for comparisons across practices. Heatmaps were generated in GraphPad Prism, Version 10 (GraphPad Software, 2023).

### Grouped trajectories

To explore patterns at the group level, participants were categorised based on the three social transition variables described above: completeness, enacted changes, and desired social transition. Sankey diagrams were then used to visualise the relationships between these categorises, showing pairwise comparisons of how many participants shared the same categorisation across each pair of variables. Specifically, three diagrams were created to illustrate shared categorisations for: (i) completeness and enacted changes, (ii) completeness and desired social transition, and (iii) enacted changes and desired social transition. Participants who shared the same category across all three variables were also identified and reported in the Results.

### Comparison of social transition group demographics

Demographic differences were assessed across the groupings for each variable, with frequencies and row-wise percentages reported for categorical variables, and means and standard deviations for continuous variables. Differences between groups were tested using Chi-square tests of independence for categorical variables and one-way ANOVAs for continuous variables. Significant differences between groups were investigated using pairwise comparisons, using proportions and their 95% confidence intervals for Chi Square results, and Tukey’s test for ANOVA results. Statistical significance was set at p<.05 for all analyses. All calculations were performed in STATA, Version 18 (StataCorp, 2023).

## Results

### Participant characteristics

There were 371 young people with survey data collected pre-MDAC appointment (i.e baseline) (mean age of 14.9 years, SD=2.2) of which most identified as binary masculine (61.1%), followed by binary feminine (18.5%), non-binary (16.6%), or unsure (3.8%) (Table 2). Further baseline demographic information about the overall sample, as well as a comparison between those subsequently included in the trajectories analysis and those lost to follow up, is reported in Table 2. No differences between the final sample and those lost to follow-up were identified.

**Table 2:** Baseline demographics for the total sample, including a split of those included in the longitudinal trajectory analysis and those lost to follow-up. Significant differences between groups are indicated by (*), where relevant.

| Demographics: | Overall<br>(n=371) | Trajectories<br>participants<br>(n=234) | Lost to attrition<br>(n=137) |
| --- | --- | --- | --- |
|  | Frequency (percentage)<br>unless specified | Frequency (percentage)<br>unless specified | Frequency (percentage)<br>unless specified |
| **Age** |  |  |  |
| **Age categories** |  |  |  |
| 8-11 years | 44 (12.4) | 33 (14.3) | 11 (8.8) |
| 12-17 years | 312 (87.6) | 198 (85.7) | 114 (91.2) |
| **Average age** |  |  |  |
| Mean (SD) | 14.9 years (SD 2.2) | 14.64 (SD 2.1) | 15.32 (SD 2.3) |
| **Country of birth, language spoken at home, socioeconomic and rural status** |  |  |  |
| **Country of birth** |  |  |  |
| Australia | 289 (84.0) | 184 (81.8) | 105 (88.2) |
| Other | 55 (16.0) | 41 (18.2) | 14 (11.8) |
| **Language spoken at home** |  |  |  |
| English | 339 (98.6) | 221 (98.2) | 118 (99.2) |
| Other | 5 (1.5) | 4 (1.8) | 1 (0.8) |
| **Socioeconomic status (IRSAD quintile)** |  |  |  |
| Quintile 1 | 40 (10.8) | 24 (10.3) | 16 (11.7) |
| Quintile 2 | 51 (13.8) | 30 (12.8) | 21 (15.3) |
| Quintile 3 | 68 (18.3) | 38 (16.2) | 30 (21.9) |
| Quintile 4 | 102 (27.5) | 72 (30.8) | 30 (21.9) |
| Quintile 5 | 110 (29.7) | 70 (29.9) | 40 (29.2) |
| **Rural status** |  |  |  |
| Metropolitan | 254 (68.5) | 167 (71.4) | 87 (63.5) |
| Regional centre | 47 (12.7) | 25 (10.7) | 22 (16.1) |
| Large rural town | 9 (2.4) | 6 (2.6) | 3 (2.2) |
| Medium rural town | 12 (3.2) | 10 (4.3) | 2 (1.5) |
| Small rural town | 49 (13.2) | 26 (11.1) | 23 (16.8) |
| **Aboriginal and/or Torres Strait Islander** |  |  |  |
| Non-Aboriginal and/or<br>Torres Strait Islander | 353 (96.2) | 224 (96.6) | 129 (95.6) |
| Aboriginal and/or Torres<br>Strait Islander | 14 (3.8) | 8 (3.5) | 6 (4.4) |
| **Gender Identity** |  |  |  |
| Binary masculine | 225 (61.1) | 148 (63.3) | 77 (57.5) |
| Binary feminine | 68 (18.5) | 41 (17.5) | 27 (20.2) |
| Non-binary | 61 (16.6) | 34 (14.5) | 27 (20.2) |
| Unsure | 14 (3.8) | 11 (4.7) | 3 (2.2) |
| **School attendance** |  |  |  |
| Attending | 312 (90.70) | 208 (92.44) | 104 (87.39) |
| Not attending | 32 (9.30) | 17 (7.56) | 15 (12.61) |

### Individual trajectories

The trajectories of social transition, organised by practice (name, pronoun, and presentation) for each participant, are shown in Figure 2. The heatmap displays the completeness metric for each practice (percentage of desired contexts where the practice has been enacted) at each wave, with participants arranged to highlight clusters of similar patterns. Visually, approximately half appear to have consistently met their social transition goals across the study period, while the remainder displays changes occurring over time.

**Figure 2:**
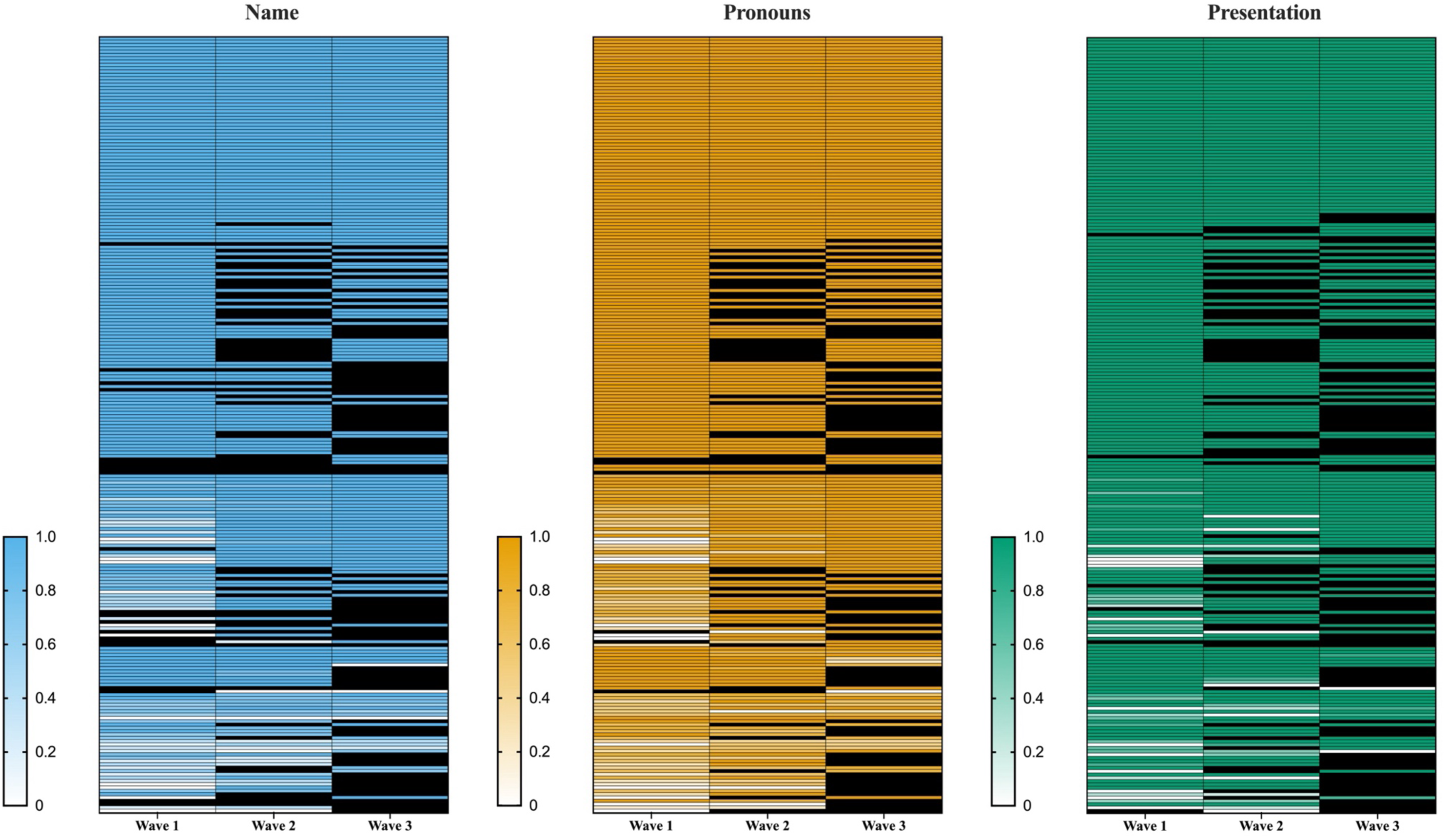
Heatmap of social transition trajectories, separated by practice. Each row represents an individual participant (n=234). Colours indicate the completeness of each practice across desired contexts (0% = white, 100% = full colour; black=missing data or no changes desired), with blue = name, orange = pronouns, and teal = presentation. Separate heatmaps are shown for each practice, with rows aligned by participant.

### Grouped trajectories

Most participants either consistently met their social transition goals across all waves (56.4%) or achieved them by the final wave (22.2%). Fewer participants were yet to meet their goals (15.4%), and a small group met them at baseline, but not by the final wave (6.0%). Regarding enacted changes, 53.8% introduced social transition practices in new contexts over the study period, while just over a quarter made no changes. A further 12.8% both added and removed contexts (in either order); only 7.7% strictly removed them. Adjustment of social transition goals were also common, with only 41.5% of the sample having stable goals. Frequency counts for all groups are presented in Table 3, with further details provided in Supplementary Table 1.

**Table 3:** Frequency of participants across categories of social transition completeness, direction of enacted changes, and changes in social transition goals (n=234)

| **Grouping variable** | **Frequency (%)** |
| --- | --- |
| Completeness of desired social transition |  |
| **Consistently meeting** social transition goals | 132 (56.4) |
| **Progression to meeting** social transition goals | 52 (22.2) |
| **Consistently not meeting** social transition goals | 36 (15.4) |
| **No longer meeting** social transition goals | 14 (6.0) |
| **Total** | **234** |
| Direction of change(s) |  |
| **Increase** in enacted changes | 123 (52.6) |
| **No change** in enacted changes | 61 (26.1) |
| **Fluctuations** (i.e. introduction and removal) | 26 (11.1) |
| **Decrease** in enacted changes (i.e. removal) | 24 (10.3) |
| **Total** | **234** |
| Adjustment of goals |  |
| **No change** in social transition goals | 92 (39.3) |
| **Increase** in social transition goals | 115 (49.1) |
| **Decrease** in social transition goals | 27 (11.5) |
| **Total** | **234** |

### Comparison of demographics by completeness metric

Comparisons of demographic variables were conducted between the groups for each variable individually (Supplementary Tables 2-4). Gender identity was significantly associated with the completeness of social transition goals (χ² (9, n=234) = 24.62, p=0.003). Post-hoc pairwise comparisons using proportions and 95% confidence intervals showed that trans boys were significantly more likely to be consistently meeting their goals (66.9%, 95%CI 58.7-74.4%) than trans girls (36.6%, 95%CI 22.1-53.1%). Non-binary young people (44.1%, 95%CI 27.2-62.1%) showed an intermediate profile that overlapped the two other groups (Supplementary Table 2). Chi-square tests also indicated associations between school attendance and both direction of enacted changes and adjustment of desires over time; however, in the pairwise comparisons, 95%CIs overlapped in all groups. Age was associated with adjustment of desires over time, with overlapping 95% confidence intervals in pairwise comparisons. No other demographic differences were observed between the groups for each variable.

### Patterns and interactions across completeness, enactment, and goals

To illustrate how participants were distributed across combinations of these variables, Sankey diagrams (Figures 3A-C) present pairwise links between categories, with band width proportional to the number of participants in each pairing. Groups sharing all three variables, along with their participant frequencies, are presented in Supplementary Table 1.

**Figure 3:**
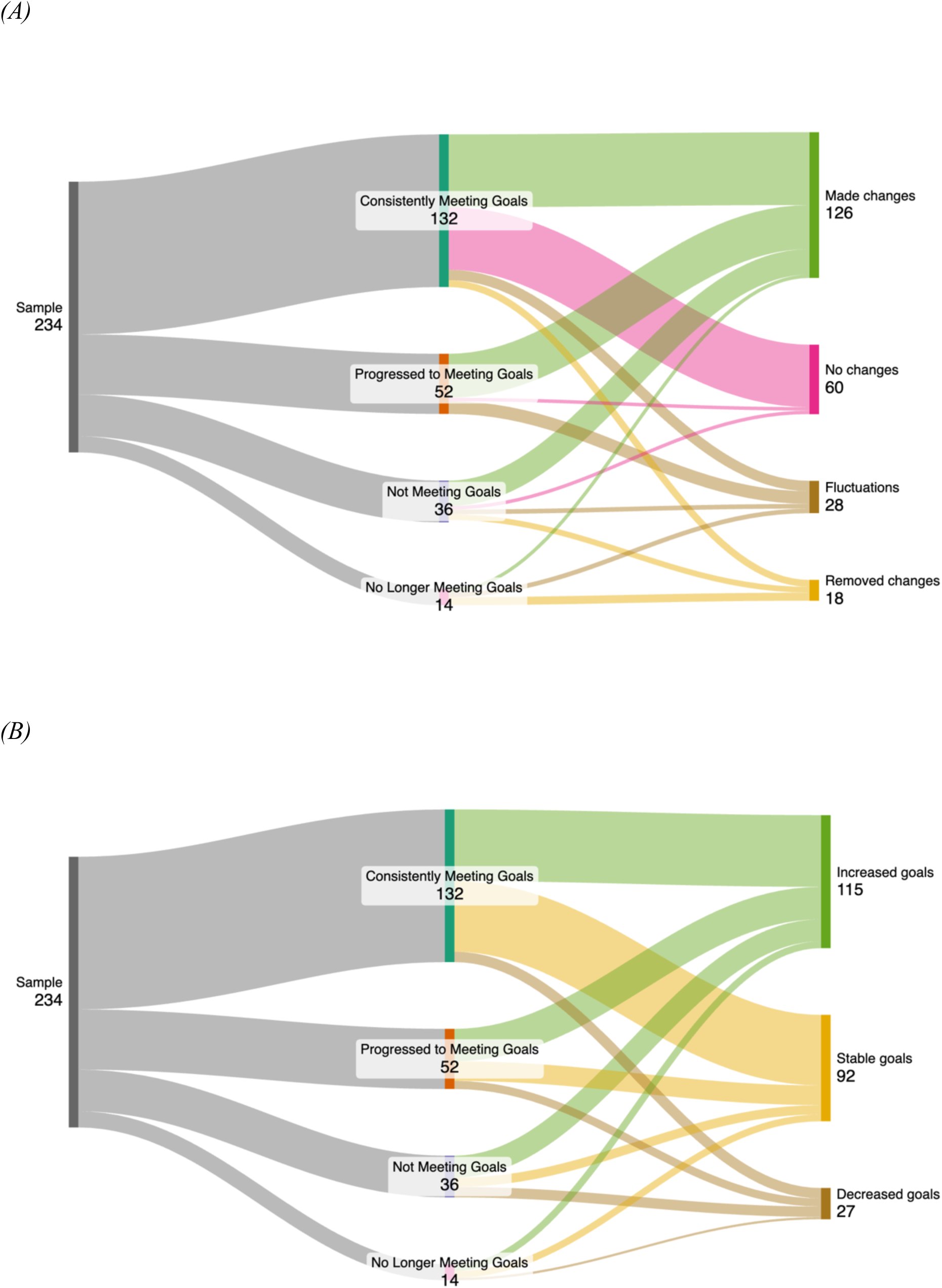
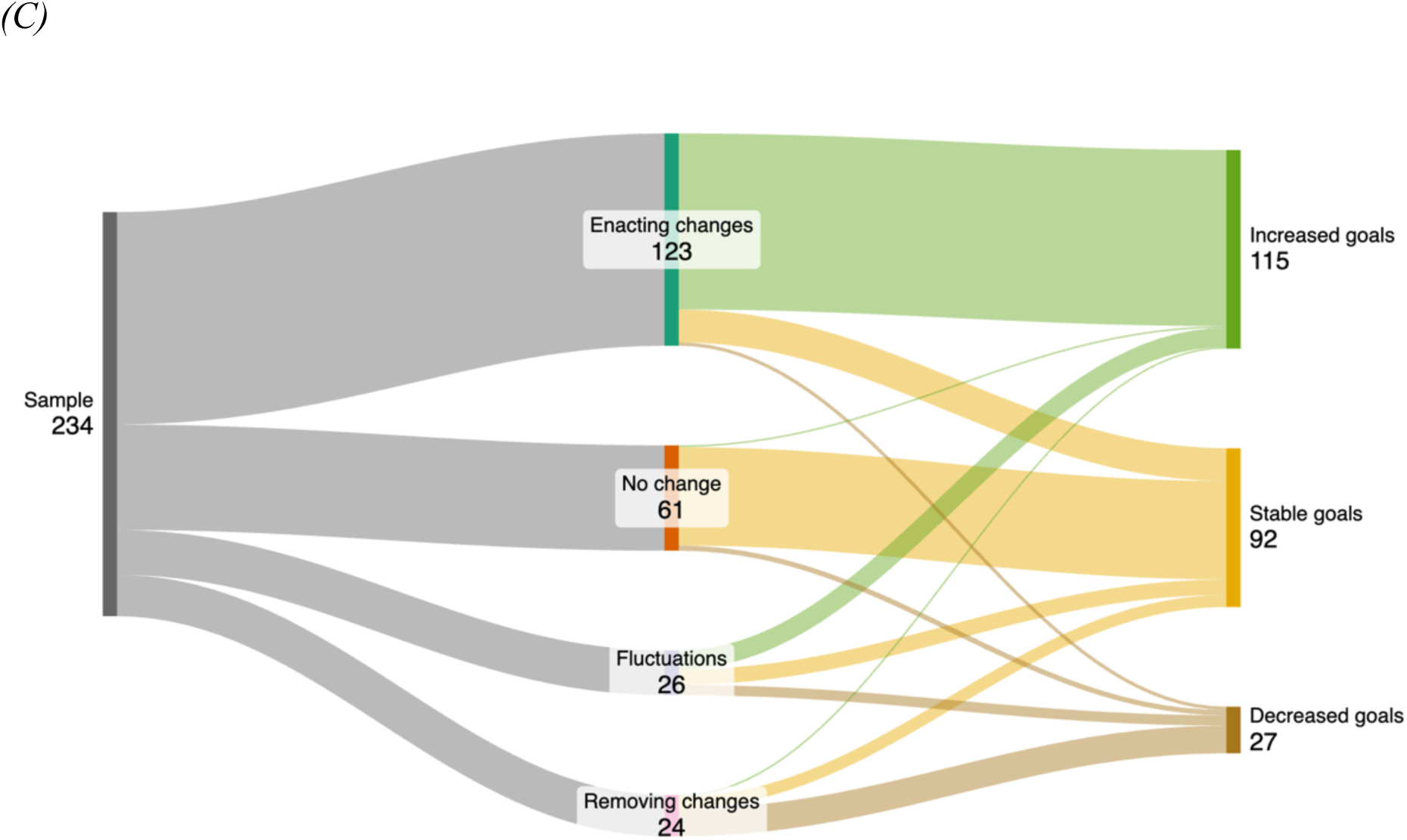
Distribution of participants with each pairwise combination of trajectory variables where band width represents the number of participants with this pairing. (A) Completeness and direction of enacted changes (B) Completeness and adjustment of goals (C) Direction of enacted changes and adjustment of goals.

#### Completeness and direction of enacted changes

Across nearly all completeness groups, the most common pattern of change was the introduction of new social transition changes (Figure 3A). Among participants consistently meeting their goals, 45.5% introduced a practice in a new context and 41.7% maintained the contexts reported at baseline. The proportion introducing practices in new contexts was higher among participants who had not yet met their social transition goals (61.1%) and those who progressed to meeting them during the study (73.1%). In contrast, among participants who initially met their goals but no longer did so at the final wave, the most frequent change was the removal of practices from previously enacted contexts (50%).

#### Completeness and adjustment of goals

Across all completeness groups, most participants either increased or maintained their social transition goals, with goal reductions being relatively uncommon (Figure 3B). In both the consistently meeting and no longer meeting groups, roughly half increased their goals and half kept them stable. Participants who progressed to meeting goals, as well as those who had not met them during the study period, were more likely to increase their social transition goals, with increases occurring approximately twice as often as stability.

#### Direction of enacted changes and adjustment of goals

Unlike the patterns observed in completeness comparisons, the direction of enacted changes and adjustment of goals varied substantially between groups (Figure 3C). Among participants not undertaking any further social transition changes, the vast majoirty maintained stable goals (93.4%). In contrast, stable goals were reported by only 15.4% of those enacting further social transition changes, with the majoirty of this group increasing their goals (82.9%). Participants who removed changes over the study period most commonly reported a decrease to their social transiton goals (66.7%); however, one third of those who removed practices did not do so in response to a change in goals. Finally, among participants reporting fluctuations in enacted changes (i.e. introductions and removals in either order), no single pattern dominated adjustment of goals, with increases, stability and decreases more evenly distrbuted (42.3%, 34.6%, and 23.1%, respectively).

## Discussion

This longitudinal analysis of social transition demonstrates the substantial heterogeneity and complexity in how young trans people desire and enact their individual social transition. While cross-sectional studies have documented this diversity in the past (Buckingham et al., 2025), following these processes over time has revealed greater nuance in these patterns. Specifically, tracking completeness in each participant’s desired social transition (i.e. goal attainment), the enactment or removal of practices, and adjustment of social transition goals illustrates how this process unfolds over time. Overall, most young people continued to enact social transition changes over the study period (63.7%), even when considered “complete” earlier. The majority adjusted their goals (60.7%), and most had met their goals by the end of the study (78.6%).

Amidst these diverse experiences, the grouping process revealed clusters of participants who, while differing in their specific changes to practices, contexts, and/or goals, followed similar overarching patterns to one another. More specifically, the completeness groups reflected whether participants were consistently meeting their social transition goals over the study period (56.4%), progressed to meeting them by the final wave (22.2%), never met these goals (15.4%), or initially met them but no longer do (6.0%). By centring completeness relative to each participant’s personal social transition goals, this strategy provides a more accurate reflection of experiences than many previous approaches. Prior studies have often operationalised social transition in more restrictive ways, such as requiring participants to present in their affirmed gender or use affirmed pronouns in all contexts (Durwood et al., 2017; Olson et al., 2016), classifying individuals using a simple ordinal scale (e.g. yes, partly, no) (Morandini et al., 2023; Sievert et al., 2020; Steensma et al., 2013), or reducing social transition status to a binary question (yes/no) (Witcomb et al., 2019). In contrast, our goal-based groupings recognise that participants may meet all their personal social transition goals without enacting every possible practice or context required by prior definitions, and they provide richer data than broad categories such as “partly”. This approach also captures adjustments to social transition goals over time, a variable overlooked in previous research, which, as highlighted earlier, occurred for the majority.

The small subset of participants who initially met their social transition goals but no longer did so by the final wave (n=14) were classified in this group for different reasons. For five participants (21.4%), this was due to an expansion of social transition goals since baseline, where they had not yet enacted all new desired changes. In contrast, nine participants had removed previously enacted changes. Among these young people, approximately half (5/9) removed only a single practice in a single context. The remaining four participants removed practices across multiple contexts; however, in each case, there was a dominant practice being removed. For example, one participant removed the use of their pronouns in all four desired contexts, while the name and presentation were only removed in a single context. Including participants’ social transition goals in the analysis prevents this group from being misinterpreted as regretful, or “desisting”; instead, these changes likely reflect responses to external pressures, such as family rejection or harassment (Turban et al., 2021b). These patterns highlight the complexity of individual experiences of social transition and underscore the need for further research to understand factors that drive the removal of previously enacted changes.

In addition to the unidirectional trajectories identified, some participants exhibited other patterns, including fluctuations or persistence with baseline social transition practices. Fluctuating patterns, characterised by introduction and removal (in either direction) of social transition changes over time, were observed in approximately one in ten participants. However, because fluctuations could only be identified in participants with three waves of data, these patterns comprised one in four participants of that subset (n=116). Fluctuating trajectories occurred in people increasing, decreasing, and persisting with baseline social transition goals, indicating that exploration can occur both in the pursuit of new goals and in navigating existing ones. They also reflect the autonomy of young people in shaping their social transition, showing that decisions about which preferences to enact, modify, or remove, are driven by existing and evolving preferences, rather than a predetermined trajectory.

Our findings indicated that trans girls were less likely than trans boys to be consistently meeting their social transition goals. While the underlying reasons for this difference were not captured in our data, trans misogyny, defined as the intersection of transphobia and misogyny experienced by trans girls and women (Arayasirikul & Wilson, 2019), may play a role. This factor has been hypothesised to be an underlying contributor to higher rates of bullying (Gower et al., 2018), less affirming school environments (Kuper et al., 2019), and older age of presentation for gender affirming care (Kahn et al., 2025) than trans girls experience. In a similar way, trans misogyny may be contributing to the lower proportion of trans girls meeting their social transition goals. No other demographic factors showed a similar association, highlighting the significance of gender identity in these outcomes. Further research is required to clarify the underlying contributors to this finding and to identify supports that may assist trans girls in navigating their social transition. These findings highlight the need for clinicians to recognise and address the distinct difficulties faced by trans girls, ensuring they are best supported to meet their social transition goals.

Several methodological features strengthened our study’s ability to capture social transition in a meaningful way. As outlined earlier, measuring multiple practices across five contexts and relative to participant-identified goals, gave the study a unique and richer perspective on social transition. Additionally, by utilising a longitudinal approach, tracking both enacted changes and desired practices over time, the grouping strategy was able to reflect meaningful patterns, rather than relying on strictly numerical algorithm-driven classifications. However, some methodological limitations may have obscured the detection of all social transition trajectories within the sample. First, many participants had already enacted social transition changes at baseline, limiting the ability to capture earlier experiences. This is particularly relevant for the group who enacted no changes and did not alter their desired practices over the course of the study (n=57), the majority of whom had already enacted all desired practices (94.7%). As such, we could not capture the patterns in which these changes occurred, nor whether any evolution of desired practices had occurred in earlier years. Second, participants with only two waves of data were treated as having a complete trajectory, which may have led to missing additional patterns and underestimating both fluctuations and the variation between participants. Third, the annual administration of the Trans20 surveys may have missed shorter-term changes in social transition, potentially underestimating fluctuations in the direction of social transition changes. Fourth, the overrepresentation of binary masculine youth in the sample (63.3%), likely reflecting the earlier presentation of assigned females for gender-affirming care (Kahn et al., 2025), may have resulted in some trajectories experienced by trans female and non-binary participants being underrepresented. Fifth, our measure of “completeness” was restricted by the practices and contexts measured in the Trans20 surveys and exclusion of “I don’t know” responses, both of which likely underestimate the full scope of social transition experiences.

Future research should build on this method of operationalising social transition (i.e. using individuals’ goals) to provide a more account of how different experiences of social transition relate to various outcomes, including gender- and mental health-related measures. Additionally, development of an improved measure of social transition through co-design with trans young people will also be important, ensuring that all meaningful changes and contexts are captured.

## Conclusion

In our longitudinal cohort of gender diverse young people, social transition encompassed diverse practices, contexts, and goals that evolved over time. Despite this diversity, distinct trajectory clusters emerged, indicating that some young people followed comparable patterns. Most participants were meeting their social transition goals at all waves, although many of these participants continued to adjust their social transition goals and make changes accordingly. Notably, goal attainment varied by gender identity, with trans boys significantly more likely to be meeting their goals than other gender groups.

## Statement of Contribution

WC led the project administration, formal analysis, writing (original draft) and visualisation; he contributed to conceptualisation and methodology. PB led validation and contributed to conceptualisation, formal analysis, methodology, supervision, visualisation, and writing (review & editing). SM contributed to supervision and writing (review & editing). KP contributed to conceptualisation, methodology, supervision, visualisation, writing (review & editing). MT led data curation as lead for the Trans20 study (Tollit et al., 2019), and contributed to conceptualisation, methodology, supervision, writing (review & editing).

## Data Sharing

The dataset generated for this study is available from the corresponding author upon reasonable request. Proposals should be directed to the corresponding author, Dr Michelle Tollit, for consideration. Data will only be shared after the proposal has been approved and once necessary ethics approvals have been obtained.

## Supplementary Material

**Supplementary Figure 1:**
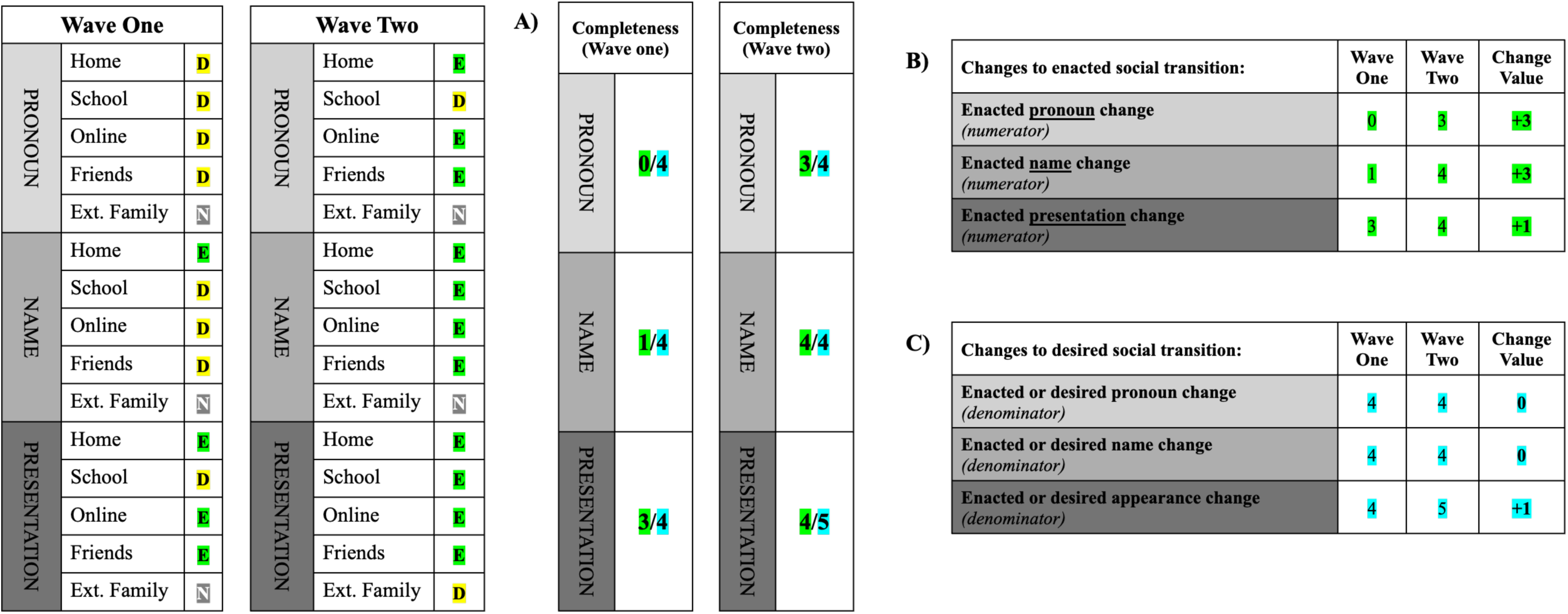
Example workflow for deriving social transition metrics. Panels show (A completeness metrics, (B) changes to enacted social transition, and (C) adjustment to social transition goals between two waves. Left-hand side data illustrate an example social transition response at Wave 1 and 2. D = desired, E = enacted, N = not desired. Values allocated green = enacted, blue = enacted or desired.

**Supplementary Table 1:**
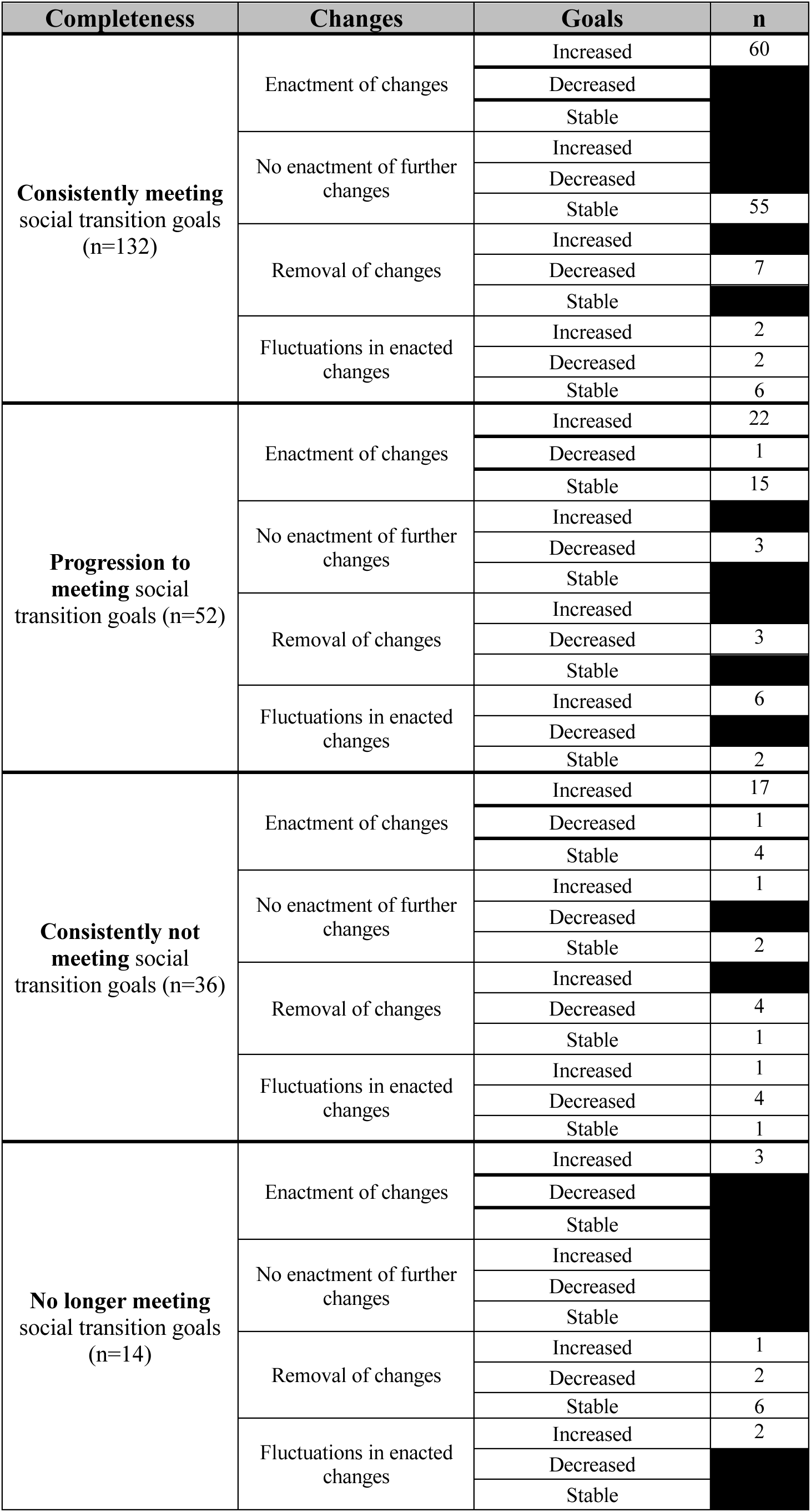
Frequency of each observed social transition trajectory, with black squares indicating a frequency count of zero.

**Supplementary Table 2:** Demographic characteristics of participants across completeness groups (consistently not meeting, consistently meeting, progression to meeting, no longer meeting goals) at baseline

| Demographics: | Consistently not meeting goals<br>(n=36, 15.4%) | Consistently meeting goals<br>(n=132, 56.4%) | Progression to meeting goals<br>(n=52, 22.2%) | No longer meeting goals<br>(n=14, 6.0%) | ANOVA/Chi Square result |
| --- | --- | --- | --- | --- | --- |
|  | Frequency (%) unless specified | Frequency (%) unless specified | Frequency (%) unless specified | Frequency (%) unless specified | p value |
| Age |  |  |  |  |  |
| Age categories |  |  |  |  |  |
| 8-11 years | 4 (12.1) | 16 (48.5) | 10 (30.3) | 3 (9.1) | 0.518 |
| 12-17 years | 31 (15.7) | 114 (57.6) | 42 (21.2) | 11 (5.6) |  |
| Average age |  |  |  |  |  |
| Mean (SD) | 14.97 (SD 1.81) | 14.70 (SD 2.03) | 14.37 (SD 2.36) | 14.29 (SD 2.37) | 0.521 |
| Country of birth, language spoken at home, socioeconomic and rural status |  |  |  |  |  |
| Country of birth |  |  |  |  |  |
| Australia (n=184) | 27 (14.7) | 104 (56.5) | 43 (23.4) | 10 (5.4) | 0.972 |
| Other (n=41) | 6 (14.6) | 23 (56.1) | 9 (22.0) | 3 (7.3) |  |
| Language spoken at home |  |  |  |  |  |
| English (n=221) | 32 (14.5) | 125 (56.6) | 51 (23.1) | 13 (5.9) | 0.904 |
| Other (n=4) | 1 (25.0) | 2 (50.0) | 1 (25.0) | 0 (0.0) |  |
| Socioeconomic status (IRSAD) |  |  |  |  |  |
| Quintile 1 | 5 (20.8) | 14 (58.3) | 3 (12.5) | 2 (8.3) | 0.358 |
| Quintile 2 | 4 (13.3) | 17 (56.7) | 6 (20.0) | 3 (10.0) |  |
| Quintile 3 | 6 (15.8) | 16 (42.1) | 13 (34.2) | 3 (7.9) |  |
| Quintile 4 | 10 (13.9) | 49 (68.1) | 10 (13.9) | 3 (4.2) |  |
| Quintile 5 | 11 (15.7) | 36 (51.43) | 20 (28.6) | 3 (4.3) |  |
| Rural status |  |  |  |  |  |
| Metropolitan | 28 (16.8) | 94 (56.3) | 37 (22.2) | 8 (4.8) | 0.800 |
| Regional centre | 3 (12.0) | 12 (48.0) | 7 (28.0) | 3 (12.0) |  |
| Large rural town | 1 (16.7) | 5 (83.3) | 0 (0.0) | 0 (0.0) |  |
| Medium rural town | 0 (0.0) | 6 (60.0) | 3 (30.0) | 1 (10.0) |  |
| Small rural town | 4 (15.4) | 15 (57.7) | 5 (19.2) | 2 (7.7) |  |
| Aboriginal and/or Torres Strait Islander |  |  |  |  |  |
| Non-Aboriginal and/or Torres Strait Islander | 33 (14.7) | 126 (56.3) | 52 (23.2) | 13 (5.8) | 0.388 |
| Aboriginal and/or Torres Strait Islander | 2 (25.0) | 5 (62.5) | 0 (0.0) | 1 (12.5) |  |
| Gender Identity |  |  |  |  |  |
| Binary masculine (n=148) | 13 (8.8) | 99 (66.9) | 27 (18.2) | 9 (6.1) | 0.003 |
| Binary feminine (n=41) | 10 (24.4) | 15 (36.6) | 14 (34.1) | 2 (4.9) |  |
| Non-binary (n=34) | 9 (26.5) | 15 (44.12) | 8 (23.5) | 2 (5.9) |  |
| Unsure (n=11) | 4 (36.4) | 3 (27.3) | 3 (27.3) | 1 (9.1) |  |
| School attendance |  |  |  |  |  |
| Attending | 29 (13.9) | 116 (55.77) | 50 (24.0) | 13 (6.3) | 0.348 |
| Not attending | 4 (23.5) | 11 (64.7) | 2 (11.8) | 0 (0.0) |  |

**Supplementary Table 3:** Demographic characteristics of participants across enactment groups (increase in changes, no changes, fluctuations in changes, and removal of previously enacted changes) at baseline

| Demographics: | Unidirectional enactment<br>(n=123, 52.6%) | No enactment of changes<br>(n=61, 26.1%) | Fluctuations in enactment<br>(n=26, 11.1%) | Unidirectional removal<br>(n=24, 10.3%) | ANOVA/Chi Square result |
| --- | --- | --- | --- | --- | --- |
|  | Frequency (%) unless specified | Frequency (%) unless specified | Frequency (%) unless specified | Frequency (%) unless specified | p value |
| Age |  |  |  |  |  |
| Age categories |  |  |  |  |  |
| 8-11 years | 21 (63.6) | 4 (12.1) | 6 (18.2) | 2 (6.1) | 0.115 |
| 12-17 years | 101 (51.0) | 56 (28.3) | 20 (10.1) | 21 (10.6) |  |
| Average age |  |  |  |  |  |
| Mean (SD) | 16.61 (SD 2.09) | 16.80 (SD 1.58) | 16.12 (SD 2.21) | 16.92 (SD 1.79) | 0.433 |
| Country of birth, language spoken at home, socioeconomic and rural status |  |  |  |  |  |
| Country of birth |  |  |  |  |  |
| Australia (n=184) | 66 (47.5) | 44 (31.7) | 14 (10.1) | 15 (10.8) | 0.104 |
| Other (n=41) | 21 (58.3) | 5 (13.9) | 7 (19.4) | 3 (8.3) |  |
| Language spoken at home |  |  |  |  |  |
| English (n=221) | 85 (49.4) | 49 (28.5) | 20 (11.6) | 18 (10.5) | 0.475 |
| Other (n=4) | 2 (66.7) | 0 (0.0) | 1 (33.3) | 0 (0.0) |  |
| Socioeconomic status (IRSAD) |  |  |  |  |  |
| Quintile 1 | 11 (45.8) | 5 (20.8) | 4 (16.7) | 4 (16.7) | 0.347 |
| Quintile 2 | 15 (50.0) | 7 (23.3) | 2 (6.7) | 6 (20.0) |  |
| Quintile 3 | 22 (57.9) | 8 (21.1) | 5 (13.2) | 3 (7.9) |  |
| Quintile 4 | 32 (44.4) | 26 (36.1) | 7 (9.7) | 7 (9.7) |  |
| Quintile 5 | 43 (61.4) | 15 (21.4) | 11.4 | 4 (5.7) |  |
| Rural status |  |  |  |  |  |
| Metropolitan | 92 (55.1) | 43 (25.8) | 16 (9.6) | 16 (9.6) | 0.341 |
| Regional centre | 11 (44.0) | 8 (32.0) | 4 (16.0) | 2 (8.0) |  |
| Large rural town | 1 (16.7) | 3 (50.0) | 2 (33.3) | 0 (0.0) |  |
| Medium rural town | 4 (40.0) | 4 (40.0) | 1 (10.0) | 1 (10.0) |  |
| Small rural town | 15 (57.7) | 3 (11.5) | 3 (11.5) | 5 (19.2) |  |
| Aboriginal and/or Torres Strait Islander |  |  |  |  |  |
| Non-Aboriginal and/or Torres Strait Islander | 120 (53.6) | 56 (25.0) | 26 (11.6) | 22 (9.8) | 0.126 |
| Aboriginal and/or Torres Strait Islander | 2 (25.0) | 4 (50.0) | 0 (0.0) | 2 (25.0) |  |
| Gender Identity |  |  |  |  |  |
| Binary masculine (n=148) | 60 (48.8) | 40 (32.5) | 14 (11.4) | 9 (7.3) | 0.132 |
| Binary feminine (n=41) | 28 (66.7) | 6 (14.3) | 3 (7.1) | 5 (11.9) |  |
| Non-binary (n=34) | 28 (50.0) | 10 (17.9) | 9 (16.1) | 9 (16.1) |  |
| Unsure (n=11) | 4 (50.0) | 3 (37.5) | 0 (0.0) | 1 (12.5) |  |
| School attendance |  |  |  |  |  |
| Attending | 76 (52.8) | 40 (27.8) | 17 (11.8) | 11 (7.6) | 0.048 |
| Not attending | 10 (33.3) | 9 (30.0) | 4 (13.3) | 7 (23.3) |  |

**Supplementary Table 4:** Demographic characteristics of participants across enactment groups (increase in changes, no changes, fluctuations in changes, and removal of previously enacted changes) at baseline

| Demographics: | No change in social transition goals (n=92, 39.3%) | Increase in social transition goals (n=115, 49.2%) | Decrease in social transition goals (n=27, 11.5%) | ANOVA/Chi Square result |
| --- | --- | --- | --- | --- |
|  | Frequency (%) unless specified | Frequency (%) unless specified | Frequency (%) unless specified | p value |
| Age |  |  |  |  |
| Age categories |  |  |  |  |
| 8-11 years | 9 (27.3) | 24 (72.7) | 0 (0.0) | 0.006 |
| 12-17 years | 81 (40.9) | 90 (45.5) | 27 (13.6) |  |
| Average age |  |  |  |  |
| Mean (SD) | 16.80 (SD 1.80) | 16.38 (SD 2.22) | 17.15 (SD 0.82) | 0.112 |
| Country of birth, language spoken at home, socioeconomic and rural status |  |  |  |  |
| Country of birth |  |  |  |  |
| Australia (n=184) | 58 (41.7) | 64 (46.0) | 17 (12.2) | 0.471 |
| Other (n=41) | 11 (30.6) | 20 (55.6) | 5 (13.9) |  |
| Language spoken at home |  |  |  |  |
| English (n=221) | 68 (39.5) | 82 (47.7) | 22 (12.8) | 0.728 |
| Other (n=4) | 1 (33.3) | 2 (66.7) | 0 (0.0) |  |
| Socioeconomic status (IRSAD) |  |  |  |  |
| Quintile 1 | 8 (33.3) | 11 (45.8) | 5 (20.8) | 0.432 |
| Quintile 2 | 13 (43.3) | 14 (46.7) | 3 (10.0) |  |
| Quintile 3 | 13 (34.2) | 20 (52.6) | 5 (13.2) |  |
| Quintile 4 | 35 (48.6) | 29 (40.3) | 8 (11.1) |  |
| Quintile 5 | 23 (32.9) | 41 (58.6) | 6 (8.6) |  |
| Rural status |  |  |  |  |
| Metropolitan | 68 (40.72) | 81 (48.5) | 18 (10.8) | 0.684 |
| Regional centre | 10 (40.0) | 12 (48.0) | 3 (12.0) |  |
| Large rural town | 4 (66.7) | 1 (16.7) | 1 (16.7) |  |
| Medium rural town | 4 (40.0) | 5 (50.0) | 1 (10.0) |  |
| Small rural town | 6 (23.1) | 16 (61.5) | 4 (15.4) |  |
| Aboriginal and/or Torres Strait Islander |  |  |  |  |
| Non-Aboriginal and/or Torres Strait Islander | 84 (37.5) | 114 (50.9) | 26 (11.6) | 0.078 |
| Aboriginal and/or Torres Strait Islander | 6 (75.0) | 1 (12.5) | 1 (12.5) |  |
| Gender Identity |  |  |  |  |
| Binary masculine (n=148) | 53 (43.1) | 59 (48.0) | 11 (8.9) | 0.606 |
| Binary feminine (n=41) | 13 (31.0) | 23 (54.8) | 6 (14.3) |  |
| Non-binary (n=34) | 21 (37.5) | 27 (48.2) | 8 (14.3) |  |
| Unsure (n=11) | 2 (25.0) | 4 (50.0) | 2 (25.0) |  |
| School attendance |  |  |  |  |
| Attending | 57 (39.6) | 73 (50.7) | 14 (9.7) | 0.027 |
| Not attending | 12 (40.0) | 10 (33.3) | 8 (26.7) |  |

